# Persistent Spike antigenemia is not associated with Post-COVID condition

**DOI:** 10.1101/2025.10.24.25338528

**Authors:** Lenny Coppens, Anna Górska, An Hotterbeekx, Lorenzo Maria Canziani, Valentin Octavian Mateescu, Elisa Gentilotti, Daniel Pirici, Matilda Berkell, Orchestra Biomarker Study Group, Surbhi Malhotra, Evelina Taconelli, Samir Kumar-Singh

**Author notes:** Department of Biomedical Engineering, Wroclaw University of Science and Technology, Wroclaw, Poland (Anna Górska). Shared senior authorship (Evelina Taconelli, Samir Kumar-Singh).

## Abstract

**Objectives:** Spike antigenemia has been proposed as one of the potential mechanisms underlying post-COVID condition (PCC). Several studies addressing Spike antigenemia have measured both free and antibody-bound Spike employing 10 mM DTT to dissociate Spike–IgG complexes.

**Methods:** Within the ORCHESTRA project, we analysed Spike and Nucleocapsid proteins in 188 participants, comprising PCC patients (n=112), non-PCC individuals (n=58) and pre-pandemic controls (n=18). For the post-pandemic cohort, 546 samples were studied from acute infection (n=99), and at 3, 6, 12, and 18 months follow-up (n=547). Impact of 10 mM DTT treatment on Spike detectability was evaluated. Correlations between Spike and proinflammatory markers were examined in both PCC and non-PCC groups.

**Results:** DTT treatment of recombinant Spike reduced its detectability in a dose-dependent manner, with a 91.3% reduction observed at 10 mM concentration (p<0.001). In plasma, DTT treatment increased detection signals in acute COVID-19 and PCC patients (n=13) by 2.6-fold, except in an acutely infected individual with exceptionally high acute Spike levels (p<0.001). Notably, pre-pandemic plasma (n=18) also exhibited a fourfold increase in measurable signal following DTT exposure (p<0.001). Analysis of free Spike and Nucleocapsid revealed that Nucleocapsid was detectable in nearly all acute cases (n=98/99), whereas Spike was observed less frequently (15/99). Neither Spike nor Nucleocapsid antigenemia differed between PCC and controls, and no association was measured between Spike levels and PCC. Interestingly, Spike levels at 3 and 6 months correlated with inflammatory cytokines IL-6, IL-2, and IL-1β, with stronger associations observed within the PCC cohort (all p<0.05).

**Conclusions:** Our findings indicate that DTT treatment induces Spike denaturation and artefactual neoepitope generation rather than genuine release of immune-complexed antigen. Furthermore, we demonstrate that PCC pathogenesis is less consistent with persistent Spike antigenemia. Together, these results call for harmonized biomarker methodologies for Spike antigenemia and more in-depth mechanistic studies of PCC.

## Introduction

Post-COVID-19 condition (PCC) is a heterogeneous syndrome marked by persistent, multisystem symptoms that persist and fluctuate months after acute infection (1, 2). The underlining mechanisms remain unclear. While initial studies showed that SARS-CoV-2 can persist in tissue for days to weeks, especially in immunocompromised individuals, others have reported the presence of viral components, such as Spike protein and viral RNA in various tissue, including blood, well beyond acute infection (3-5). These findings, along with reports of elevated Spike in PCC patients, support the hypothesis that Spike may contribute to PCC pathogenesis (6-8). However, not all studies support this view (9-11).

Several studies reporting Spike in PCC have employed dithiothreitol (DTT), which disrupts disulfide bonds to dissociate antigen-antibody complexes, thereby enabling quantification of both antibody-bound and free Spike (6-8). However, the biological relevance of antibody-bound Spike and the interpretability of DTT-treated measurements remain unresolved. The pathogenic potential of such complexes is poorly defined. Fully glycosylated Spike and S1 domain have molecular weights of ∼180–200 kDa and ∼90–120 kDa, respectively, compared with ∼150 kDa for intact IgG (12). Given inter-individual variability in HLA alleles and B- and T-cell receptor repertoires, the epitopes targeted by anti-Spike antibodies are expected to generate highly heterogenous Spike-IgG immune complexes, making it difficult to envisage a consistent, biologically relevant gain-of-function mechanism contributing to PCC.

In addition, DTT itself may directly alter Spike structure. Prior studies have shown that low millimolar DTT concentrations, even below those used in current assays (6-8, 13), disrupt Spike receptor-binding domain (RBD), which contains three disulfide bonds, to an extent that inhibits viral replication (14). These considerations raise fundamental concerns about whether DTT-based assays truly reflect circulating or biologically relevant antigen levels.

To address this, we analysed plasma from well-characterized participants in the prospective ORCHESTRA cohort, followed for up to 18 months after initial SARS-CoV-2 infection, to investigate the presence of Spike or Nucleocapsid antigenemia. In parallel, we systematically evaluated the impact of DTT on Spike detection.

## Material and Methods

### Study population

Participants with RT-PCR-confirmed SARS-CoV-2 infection were prospectively enrolled at the University Hospital of Verona within the ORCHESTRA project. This sub-study cohort included 112 individuals meeting WHO criteria for PCC and 58 age- and sex-matched non-PCC control individuals (**SI Table 1**). Peripheral blood was collected during acute infection (n=99) and at 3, 6, 12, and 18 months post-infection (n=447; **SI Table 2**). Pre-pandemic controls comprised 18 surgical patients. In total, 564 samples were analyzed. All participants provided written informed consent, and the study was approved by the Verona Ethics Board in accordance with the Declaration of Helsinki.

### Antigen, serological, and cytokine assays

To assess Spike protein stability and recovery, plasma samples (pre-pandemic and pandemic), and recombinant Spike were treated with freshly prepared dithiothreitol (DTT) containing protease inhibitors and EDTA, incubated at 37°C for 15 minutes, and diluted to 1 mM DTT, as previously described (6, 13). Free Spike and Nucleocapsid antigens as well as DTT-treated plasma samples were quantified using ultrasensitive S-PLEX SARS-CoV-2 assays (Meso Scale Discovery), with LLOQs of 350 fg/mL (S) and 172 fg/mL (N). Serological responses to Spike and Nucleocapsid, along with select cytokines were measured using MSD platforms (see Supplementary Information).

### Statistics

Non-normal data were log_10_-transformed or analysed with non-parametric methods. Associations with PCC or symptom clusters were evaluated using multivariate mixed-effects logistic-regression models, and correlations between Spike antigen, serology, and cytokines were assessed using Pearson’s or Spearman methods. Detailed descriptions of statistical procedures, model specifications, and additional analyses are provided in the Supplementary Information.

## Results

### Low molar concentrations of DTT denature SARS-CoV-2 Spike protein

To examine the effect of DTT on Spike protein, recombinant mammalian-expressed Spike was incubated with increasing concentrations of DTT (5, 10, 15, and 50 mM) and compared to untreated controls. Quantitative immunoassays showed a dose-dependent reduction in detectable Spike, with significant loss at 5 mM and near-complete ablation at 50 mM. At 10 mM DTT, used in prior studies (6, 7), the signal decreased by 91.3 ± 0.1% across two independent experiments in triplicate (*p*<0.001, **Fig. 1A; SI Fig. 1B**). Because plasma components can interfere with DTT activity, we next spiked recombinant Spike into pre-pandemic plasma (n=6) and observed a similar 94.95% reduction in Spike detectability (*p*<0.001; **Fig. 1B**). To extend these findings in a biological context, we analysed pandemic plasma treated with DTT (n=13). DTT increased antigen signal 2.6-fold on average, except in one acute case with exceptionally high Spike levels where it decreased by 97% (*p*<0.001 **Fig. 1C**). Pre-pandemic plasma (n=6) treated the same way showed a fourfold increase (*p*<0.001 **Fig. 1D**), indicating that DTT denatures Spike and induces artefactual neoepitope formation from unrelated plasma proteins rather than specifically releasing Spike from immune complexes.

**Figure 1:**
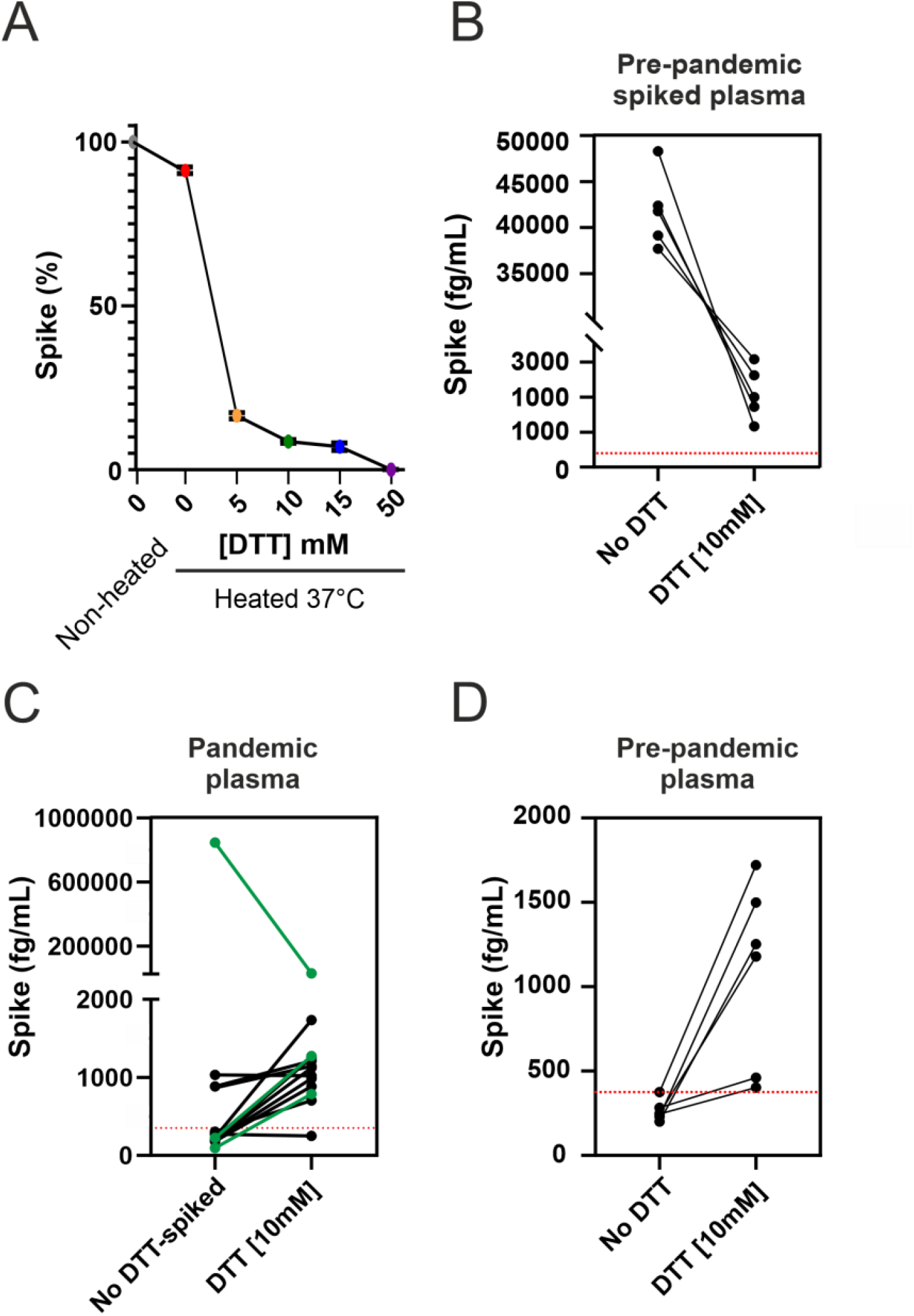
Effect of DTT treatment on spike protein signal. (**A**) DTT treatment induces a dose-dependent decrease in recombinant spike protein signal. Data are representative of two independent experiments; each point represents the mean of triplicate measurements, and error bars denote the standard deviation. (see also S.I. Figure 1). (**B**) Spiking recombinant Spike protein in pre-pandemic plasma shows a significant decrease in Spike concentration after standard 10 mM DTT-treatment (**C**) Post-pandemic samples generally exhibit significant increase in Spike signal following standard 10 mM DTT treatment, except for one sample showing a decrease; acute-phase samples are indicated in green. (**D**) All pre-pandemic samples display an increase in Spike signal after standard DTT treatment. The red line denotes the lower limit of quantification (LLOQ).

### Free Spike protein is detectable up to 18 months but not elevated in PCC

We next quantified free Spike and Nucleocapsid in plasma from 188 individuals, including 170 SARS-CoV-2–infected participants and 18 pre-pandemic controls. All pre-pandemic samples were negative for both antigens, except for 1 marginal Spike reading, confirming assay specificity. In acute infection, Nucleocapsid was quantifiable in nearly all cases (98/99), often exceeding 10^6^ fg/mL whereas Spike antigenemia was less frequent (15/99) but reached up to 10^5^ fg/mL in some participants (**Fig. 2A**). Hospitalized individuals with quantifiable Spike had higher levels of both antigens (*p*<0.01) and displayed elevated anti-Nucleocapsid IgG alongside reduced anti-Spike IgG (*p*<0.01; **SI Fig. 2A**).

**Figure 2:**
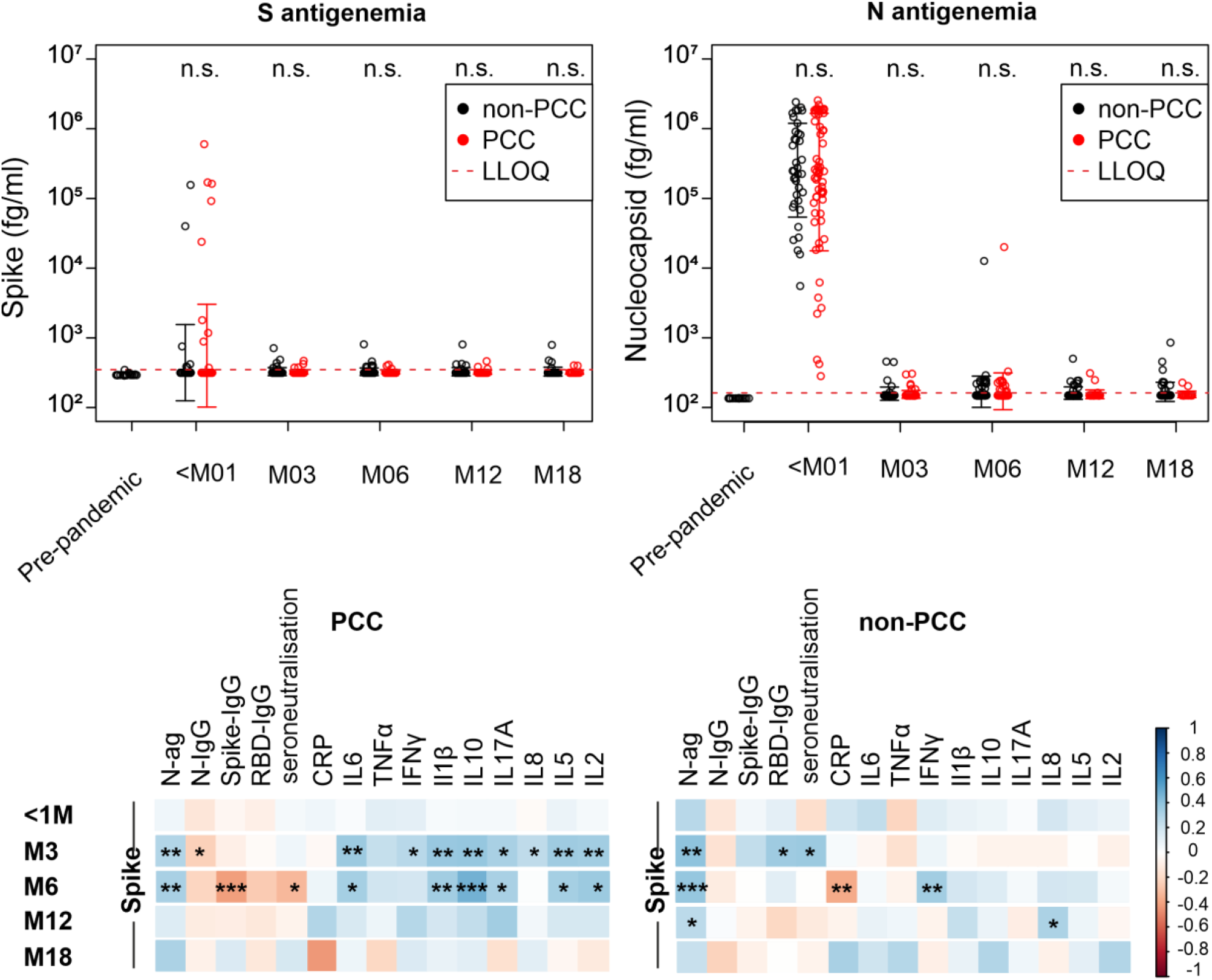
Longitudinal dynamics of Spike and Nucleocapsid antigens and association of spike with cytokines. (**A**) Longitudinal analysis showing elevated Spike and Nucleocapsid antigen levels during the acute COVID-19, with a decline observed in both fully convalescent and post-COVID condition (PCC) patients in the post-acute phase. Each point represents the mean value, and error bars denote the standard deviation. (**B**) Spearman correlation heatmaps of S antigens with Nucleocapsid antigen, anti-Nucleocapsid and anti-Spike IgGs, and cytokines at acute (<1M) and post-acute phases, stratified by PCC status (see also SI Figure 3 for analysis across all patients). Asterisks indicate statistical significance of unadjusted p-values: *p < 0.05, **p < 0.01, ***p < 0.001. Only correlations involving Spike antigen are shown.

At 3, 6, 12, and 18 months post-infection, low level antigen persistence was observed in a subset of participants, though concentrations were substantially lower than during the acute phase. Persistent antigenemia, defined as detectable Spike or Nucleocapsid antigens beyond the acute phase, did not differ between individuals with or without PCC. Multivariate mixed-effects logistic-regression, adjusting for age, sex, timepoint, and repeated measures, confirmed no association of Spike (OR=0.45, *p*=0.14) or Nucleocapsid (OR=0.61, *p*=0.28) with PCC or its symptom clusters (2). By contrast, the likelihood of PCC declined progressively over time, with significantly lower odds at 6, 12, and 18 months compared with 3 months post-infection (all *p*<0.05; **SI Table 2**).

### Associations between circulating Spike and inflammatory cytokines

Spike antigen correlated with antibody and cytokine responses most strongly at 3–6 months post-infection, particularly with anti-Nucleocapsid IgG, Nucleocapsid antigens and pro-inflammatory cytokines (IL-6, IL-2, IL-1β; **SI Fig. 3**). Stratification by PCC revealed distinct patterns: in PCC patients, Spike showed consistent positive correlations with both humoral and inflammatory markers, whereas non-PCC individuals exhibited fewer associations, consistent with more resolved immune responses (**Fig. 2B**). These findings suggest that circulating Spike is linked to higher immune activation early in convalescence, with associations diminishing as antigen levels decline and immune memory matures.

## Discussion

Previous reports of persistent Spike in long COVID often relied on 10 mM DTT to dissociate antigen–antibody complexes, enabling detection of both free and antibody-bound Spike (6-8). We show that DTT treatment denatures both native and recombinant Spike in a dose-dependent manner and increases signal even in pre-pandemic plasma, raising concerns about assay specificity when DTT is used.

Longitudinal analysis up to 18 months revealed low-level Spike and Nucleocapsid persistence in a subset of individuals, but neither was associated with PCC presence or ORCHESTRA-define symptom clusters (2). Correlations between Spike and inflammatory cytokines were confined to PCC patients, suggesting that minimal antigen persistence may still drive immune dysregulation in susceptible hosts. Nevertheless, these results argue against widespread viral persistence as the primary driver of PCC and challenge the validity of circulating Spike as a biomarker (6-8) for this syndrome.

This study has limitations. Assay antibodies may vary in affinity across viral variants, and antigen levels comparable to those observed here and elsewhere (6-8) can also occur immediately post-vaccination (15). Future work should therefore account for variant-specific reactivity and vaccination status.

In conclusion, our findings have several implications for ongoing PCC research and clinical practice. First, they caution against over-interpreting persistent viral antigenemia as a biomarker or therapeutic target for PCC without further validation. Second, they highlight the need for methodological rigor and standardization in future PCC biomarker studies, particularly regarding sample preparation and antigen detection protocols. These results underscore the importance of investigating the mechanisms underlying post-acute infection syndromes mechanisms across diverse pathogens, as they can follow multiple different infections yet exhibit shared clinical features. Such cross-pathogen studies will be essential for strengthening pandemic preparedness (1). Finally, our results highlight the need for a deeper understanding of how Spike antigenemia might be linked to immune dysregulation-driven PCC pathogenesis.

## Data Availability

All data produced in the present study are available upon reasonable request to the authors

## Declaration of Competing Interest

The authors declare that there are no competing financial interests or personal relationships that could have been perceived as influencing the research presented in this manuscript.

## Author contribution

Conceptualization overall study supervision: SK-S; Clinical lead: ET; Clinical data and sample collection: EG, ET, LMC; Database collation: AG, MB, AH, LMC, and LC; Laboratory analysis: LC, AH, and VOM; Data interpretation: SK-S, LC, AH, DP, SM, and ET; First draft LC and SK-S. All authors read, gave input, and approved the final manuscript.

## Ethics

Participants were recruited from the Infectious Diseases Section of the University Hospital of Verona from February 10, 2021 to October 17, 2023. All volunteers provided informed, written consent before study participation. This study was approved by the University Hospital Verona Ethics Board (protocol number: 19293) and conducted in accordance with the Declaration of Helsinki.

## Funding Statement

This work is supported by the ORCHESTRA project, which has received funding from the European Union Horizon 2020 research and innovation program under grant agreement No 101016167.

## Supplementary material

### METHODS

#### 1.1.1 Overall Design

This study examined individuals from the acute phase of SARS-CoV-2 infection through 18 months post-infection to investigate differences in unbound SARS-CoV-2 plasma antigen levels between participants with and without Post-COVID condition (PCC). Additionally, the impact of DTT on SARS-CoV-2 Spike detection was assessed.

#### 1.1.2 Participants

We studied two groups of participants who were recruited from the Infectious Diseases Section of the University Hospital of Verona as part of the prospective, multicenter ORCHESTRA study. The overall Verona cohort within ORCHESTRA is larger, and the selection of participants has been described previously (2). Briefly, patients requiring hospital admission during the acute phase of infection as well as outpatients were eligible for enrolment. Follow-up assessments were conducted at 3, 6, 12 and 18 months after the initial infection. For the present study, we analyzed a sub-cohort of 188 participants, including 112 patients with symptoms consistent with PCC, 58 COVID-19 convalescent controls who were matched for age and sex and 18 pre-pandemic patients. All pandemic patients had a laboratory-confirmed SARS-CoV-2 infection during the first sampling time point. PCC was defined as self-reporting at least one of the following symptoms: arthralgia, cough, dyspnea, fatigue, fever, memory loss, myalgia, loss of taste or smell, at any time in the follow-up. PCC cases were further classified into clinical phenotypes established by ORCHESTRA (2). All participants provided written informed consent before participating in the study. The study protocol was approved by the Ethics Board of the University Hospital of Verona and was conducted in accordance with the principles of the Declaration of Helsinki. Samples were stored in the Biobank of Antwerp University Hospital at −80°C until analysis.

#### 1.1.3 Characteristics of the Study Cohort

A sub-cohort of 170 individuals from the Verona site of the ORCHESTRA H2020 project was analysed to investigate the potential role of post-COVID antigenemia in the development of PCC. The cohort included 112 participants diagnosed with PCC (62 females, 50 males) and 58 control subjects (29 females, 29 males) with no reported history of PCC. The mean age was comparable between groups, with 59.5 ± 14.9 years in the PCC group and 59.3 ± 13.1 years in the control group. Hospitalization during the acute phase of COVID-19 was more frequent among individuals with PCC (33% vs. 15%; *p*=0.018), and moderate to severe disease, as defined by a WHO progression scale score of 5–8, was significantly more prevalent in this group (37% vs. 16%; *p*=0.023). The proportion of individuals who received monoclonal antibody treatment did not differ significantly between groups (**SI Table 1**).

#### 1.1.4 Antigen Measurement

Peripheral blood was collected in EDTA-coated tubes and plasma was stored at −80^°^C until further analyses. Once-thawed plasma was analysed for SARS-CoV-2 Spike and Nucleocapsid (N) proteins using MSD ultrasensitive S-plex assays (K150ADJS and K150ADHS). For DTT treatments, a fresh 1 M dithiothreitol (DTT) solution was prepared in presence of protease inhibitors and EDTA (Halt™ Protease and Phosphatase Inhibitor Cocktail, Thermo Fisher). Recombinant Spike, pre-pandemic, and pandemic plasma samples were incubated under conditions replicating those previously described (6, 13). Assay diluent was added post-incubation to adjust the final DTT concentration to ∼1 mM.

Quantitation was performed against a recombinant antigen standard curve fitted to a four-parameter logistic (4PL) model. The LLOQ of the assays was defined as the lowest concentration yielding coefficients of variances (CVs) of less than 25%. The lower limit of detection (LLOD) was 95 fg/mL for Spike and 62 fg/mL for Nucleocapsid; however, values below the LLOQ (Spike, 350 fg/mL; Nucleocapsid, 172 fg/mL) are not considered reliably quantifiable and were not taken into account. The Nucleocapsid antigen assay uses recombinant full-length Nucleocapsid protein as a standard, and monoclonal capture/detection antibody generated by MSD against the full-length recombinant Nucleocapsid protein. The Spike antigen assay utilizes recombinant receptor binding domain (RBD) of the Spike subunit as a standard, and monoclonal capture/detection antibody generated by MSD against recombinant RBD.

For serological assays peripheral blood was collected in 10 mL serum tubes (Vacutainer K2E, BD Biosciences) and samples were prepared within 3 hours of blood collection. Seroneutralizing capacity, anti-Nucleocapsid, anti-Spike, and anti-Receptor-binding domain (RBD) SARS-CoV-2 IgG titers were measured in serum samples using a multiplexed panel 2 (Meso Scale Discovery, K15383U) and data provided in WHO-recommended BAU units. Plasma levels of cytokines, chemokines and growth factors (CCGs) were quantified using the U-plex assay from MSD, according to the manufacturer instructions. The analytes included C-reactive protein (CRP), interleukin (IL)-1β, IL-6, IL-8, IL-10, IL-17A, interferon-γ (IFN-γ), and tumor necrosis factor-α (TNF-α). Measurements were performed in randomized batches.

#### 1.1.5 Statistical analysis

For the comparison of antigen levels across the study groups, five distinct time periods were defined relative to the onset of acute COVID-19 symptoms: 2 weeks (±2 weeks), 3 months (±1 month), 6 months (±1 month), 12 months (±1 month), and 18 months (±1 month). All statistical analyses and visualizations were performed using Microsoft Excel, SPSS v27 and R Studio (R version 4.4.3). Normality of the concentration distribution was assessed using the Shapiro–Wilk test. For non-normally distributed variables, either parametric analyses were performed on log_10_-transformed values or non-parametric methods were applied as appropriate. Paired comparisons were conducted using paired t-tests when parametric assumptions were met; otherwise, the non-parametric equivalent was used. Associations between PCC or its symptom clusters and measured variables were evaluated using linear mixed-effects models adjusted for repeated measures, time point, age, and gender. Correlation analyses were limited to Spike antigen and its relationship with serology and cytokine data. These correlations were first evaluated in the full cohort and subsequently stratified by PCC versus non-PCC. Pearson’s correlation was used for normally distributed data, and Spearman’s rank correlation for non-normal data. CRP was analysed using a clinically relevant cutoff of <5mg/L. Statistical significance in the figures is indicated as follows: one, two, or three stars correspond to p < 0.05, p < 0.01, and p < 0.001, respectively.

## Supplementary Table

**SI Table 1:** Demographic details of the studied cohort.

| Variables | Non-PCC control<br>(n=58) | PCC<br>(n=112) | p-value |
| --- | --- | --- | --- |
| Age (mean (SD)) | 59.48 (14.88) | 59.28<br>(13.13) | 0.926 |
| Gender (%) |  |  |  |
| male | 29 (50.0) | 50 (44.6) | 0.616 |
| female | 29 (50.0) | 62 (55.4) |  |
| Hospitalization<sup>a</sup> (%) | 9 (15.5) | 38 (33.9) | 0.018 |
| mAb treatment<sup>a</sup> (%) | 34 (58.6) | 55 (49.1) | 0.31 |
| WHO severity<sup>a</sup> (%) |  |  |  |
| 1-5 | 47 (83.9) | 71 (65.7) | 0.023 |
| 6-8 | 9 (16.1) | 37 (34.3) |  |

<sup>a</sup>at acute COVID-19

**S.I. Table 2:** Sample analysis across study timepoints. The numbers indicate participants with available plasma samples analysed at each visit.

| **Group** | **<M01**<br>**(acute)** | **M03** | **M06** | **M12** | **M18** | **Pre-**<br>**pandemic** |
| --- | --- | --- | --- | --- | --- | --- |
| **PCC (n=112)** | 61 | 77 | 68 | 37 | 22 | – |
| **Non-PCC control (n=58)** | 38 | 56 | 79 | 56 | 52 | – |
| **Pre-pandemic (n=18)** | – | – | – | – | – | 18 |
| **Total (n=188 patients; 564 plasma)** | 99 | 133 | 147 | 93 | 74 | 18 |

**S.I. Table 3:** Multivariate logistic regression models for general PCC and the four ORCHESTRA-defined symptom clusters (C1: chronic fatigue-like; C2: respiratory; C3: chronic pain; C4: neurosensory, Ref. (2), Main text). Patient-level clustering was accounted via a random intercept. Significance levels: *p < 0.05; **p < 0.01; ***p < 0.001.

|  | PCS |  | C1 |  | C2 |  | C3 |  | C4 |  |
| --- | --- | --- | --- | --- | --- | --- | --- | --- | --- | --- |
|  | OR | Sig. | OR | Sig. | OR | Sig. | OR | Sig. | OR | Sig. |
| **S antigen** | 0.451 | ns | 0.465 | ns | 1.442 | ns | 1.785 | ns | 0.451 | ns |
| **N antigen** | 0.611 | ns | 0.971 | ns | 0.168 | ns | 0.622 | ns | 0.611 | ns |
| **Female sex** | 1.556 | ns | 0.622 | ns | 2.602 | ns | 0.316 | ns | 1.556 | ns |
| **Age** | 0.694 | ns | 1.025 | ns | 0.954 | ns | 0.986 | ns | 0.991 | ns |
| **Month 6** | 0.483 | ** | 0.414 | ns | 0.436 | ns | 0.376 | ns | 0.483 | ns |
| **Month 12** | 0.351 | *** | 0.895 | ns | 0.035 | * | 0.287 | ns | 0.351 | * |
| **Month 18** | 0.089 | *** | 0.036 | *** | 0.026 | * | 0.012 | *** | 0.089 | *** |

## Supplementary Figures

**SI Figure 1:**
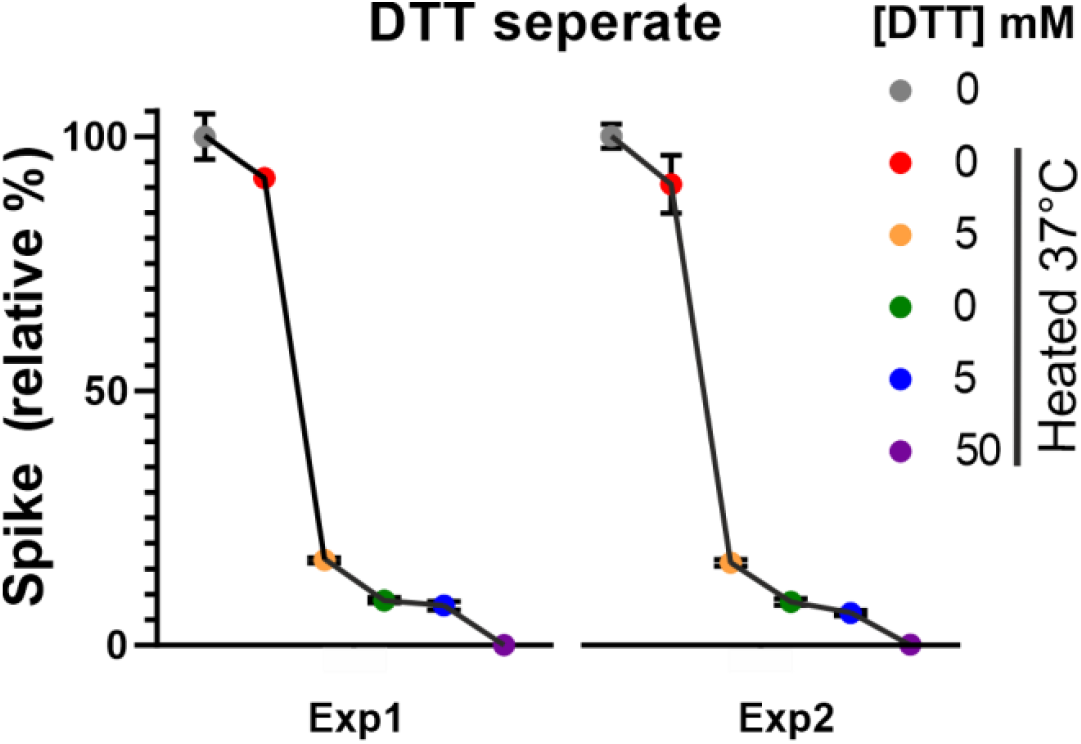
Effect of DTT on Spike detectability in two independent experiments. DTT treatment induces a dose-dependent decrease in Spike protein signal. Graph shows two independent experiments (Exp), with each point representing the average of triplicate measurements.

**SI Figure 2:**
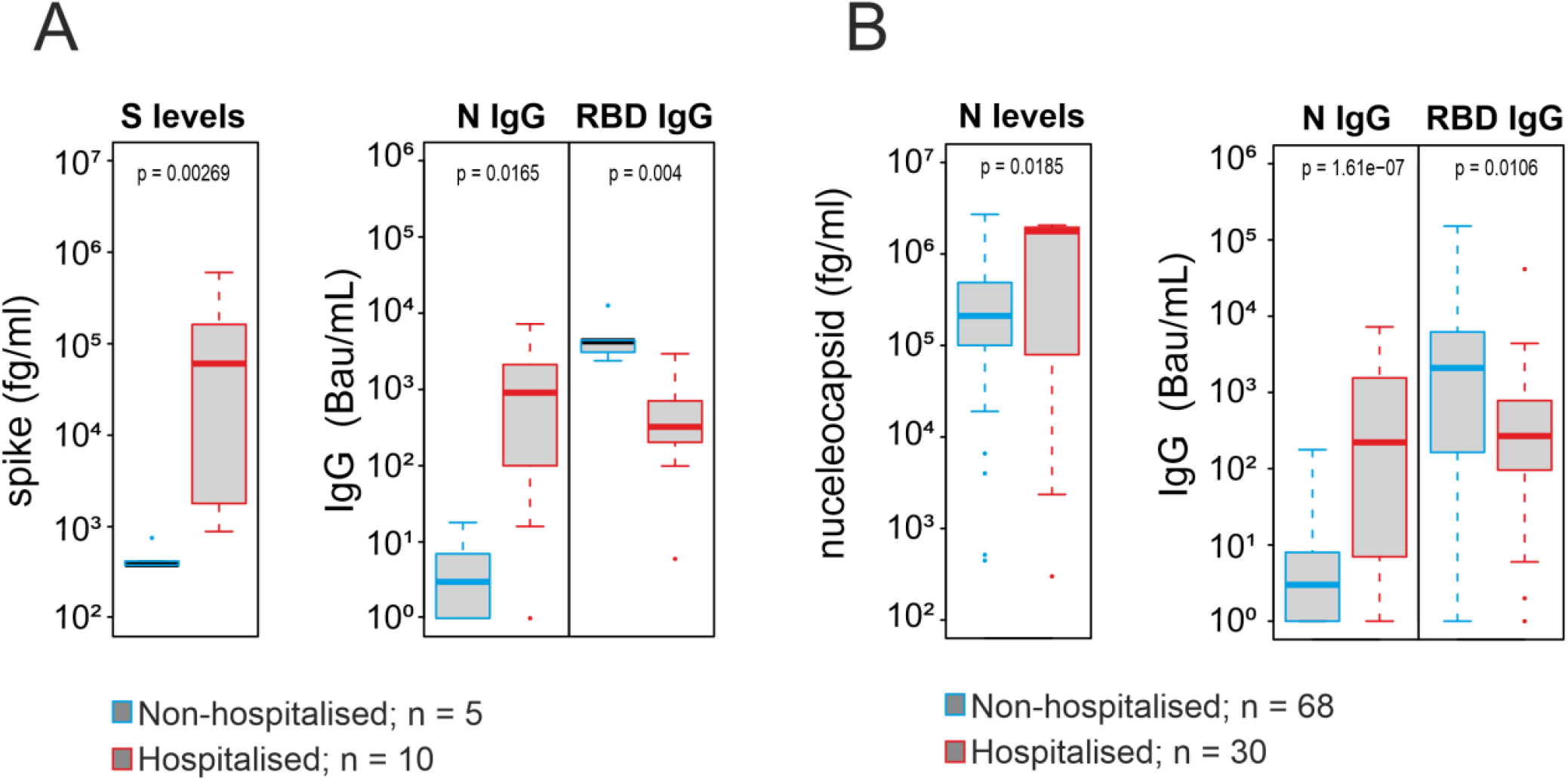
Antigen and antibody profiles in hospitalized patients. **(A)** Hospitalized patients with quantifiable Spike antigen showed elevated detectable Spike antigen and anti-Nucleocapsid IgG antibodies but reduced anti-RBD antibody levels. **(B)** Hospitalized patients with quantifiable Nucleocapsid antigen showed elevated detectable Nucleocapsid antigen and anti-Nucleocapsid IgG antibodies but reduced anti-RBD antibody levels.

**SI Figure 3:**
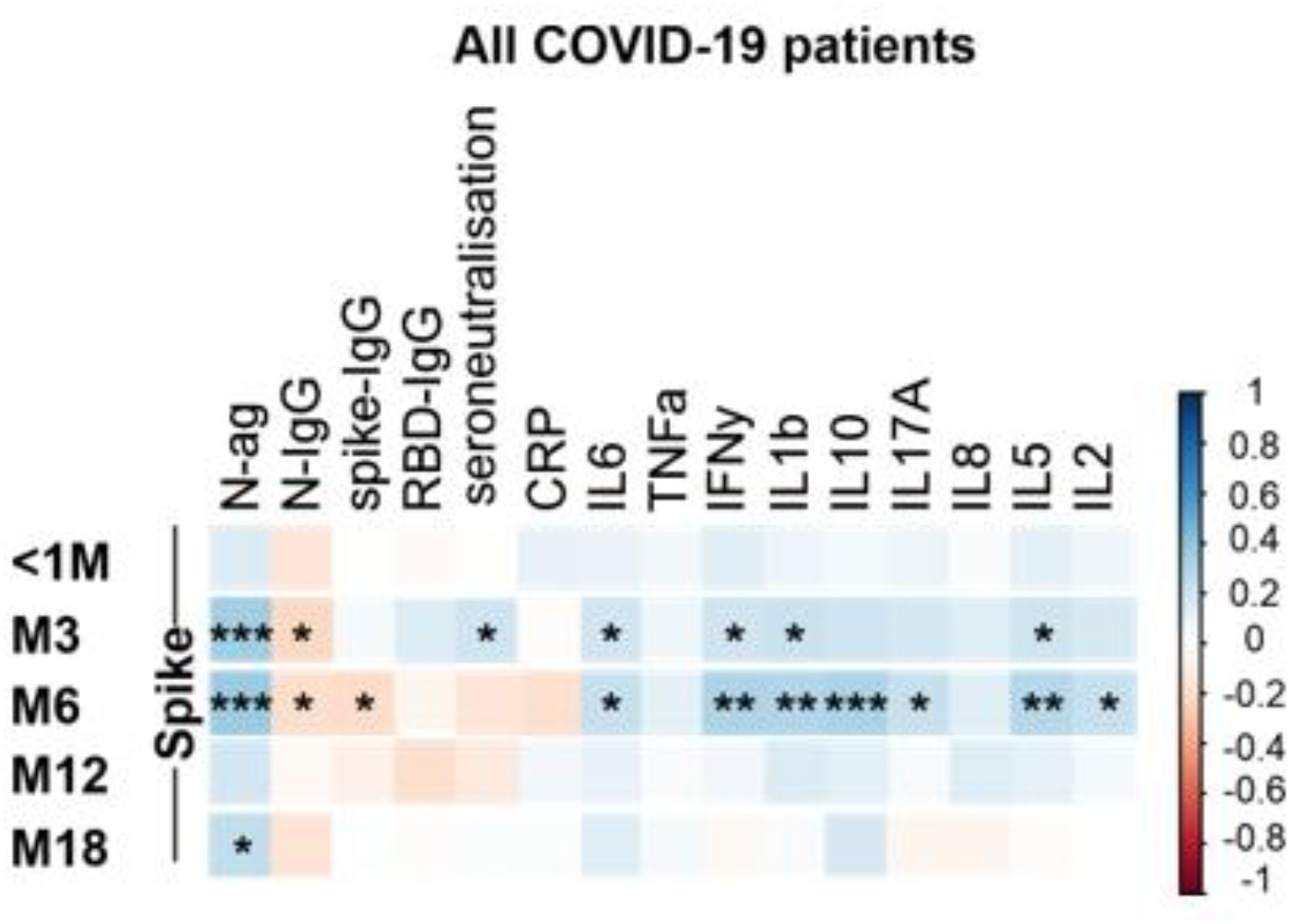
Correlations between Spike antigen, anti-Nucleocapsid, anti-Spike, and cytokines across all patients. Heatmaps show Spearman correlations between Spike antigens and various cytokines at baseline (<1M) and post-acute time points. Asterisks indicate statistical significance of unadjusted p-values: \**p* < 0.05, \*\**p* < 0.01, \*\*\**p* < 0. 001. Only correlations involving Spike antigens are displayed.

## Notes

**Funding**: This work is supported by the ORCHESTRA project, which has received funding from the European Union’s Horizon 2020 research and innovation program under grant agreement No 101016167

**Conflicts of interest**: The authors declare no conflict of interest

### Competing Interest Statement

The authors have declared no competing interest.

### Author Declarations

This study was approved by the University Hospital Verona Ethics Board (protocol number: 19293) and conducted in accordance with the Declaration of Helsinki.

